# Phase II Trial Evaluating the Association of Peripheral Blood Immunologic Response to Therapeutic Response After Adjuvant Treatment with Immune Checkpoint Inhibition (ICI) in Patients with Newly Diagnosed Glioblastoma or Gliosarcoma

**DOI:** 10.64898/2025.12.23.25342908

**Authors:** Neuro-Oncology Clinical Research Program, Kevin Camphausen

## Abstract

**Background:** Glioblastoma (GBM) represents an aggressive malignancy with limited therapeutic options. The immunosuppressive nature of GBM may be reversible with immune checkpoint inhibitor (ICI) treatment, however, initial studies have yet to demonstrate this. It is postulated that trafficking of peripherally activated lymphocytes may play a role in generating a robust intracranial immune response. Therefore, a blood-based assay to identify peripheral blood response may both predict response and better identify the ideal patient populations for future ICI clinical trials.

**Methods:** This was an open-label, Phase II, investigator-initiated exploratory study of patients with newly diagnosed GBM who completed maximal tumor resection and concurrent chemoradiation followed by standard adjuvant temozolomide and the combination of Nivolumab and Ipilimumab. The primary objective was to determine if the outcome, as measured by overall survival, is improved in patients when treatment with immune checkpoint inhibitors results in an immune response in peripheral blood T lymphocytes. The immune response is defined as changes in the CD4^+^/CD8^+^ precursor frequency and expansion index compared to the overall survival (OS) measured in months.

**Results:** The study closed to enrollment early due to a shift in clinical priorities after the accrual of 40 patients. Twenty-three patients have died of their disease, and adequate samples for the primary analysis were available for 17 of these patients. The median OS for the 17 patients was 19 months (range 9-45months). For the four immune measurements, patients were categorized as *reactive, indeterminate*, or *suppressed* based on pre-defined protocol criteria. Only two patients were classified as *reactive* across all four measurements, and their median OS was 17.5 months, compared with 21 months for patients classified as suppressed. Across each of the 4 individual immune measurements, no statistical difference in OS were observed between *reactive* and *suppressed* groups.

**Conclusion:** In this limited cohort, no detectable difference in the OS was observed between patients with a *reactive* immune signature and those with a *suppressed* immune signature.

## INTRODUCTION

Malignant gliomas, in particular glioblastoma (GBM), are almost always fatal despite aggressive treatment, including surgery, radiation, and chemotherapy with temozolomide (TMZ)(1). Though numerous trials have attempted to improve outcomes by modifying treatment regimens or adding novel agents, none have significantly improved survival. Recent efforts have focused on immunotherapy, including tumor vaccines (e.g., CDX-110, ICT-107, HSPPC-96), and checkpoint inhibitors, though promising early results have also not translated into success in Phase 3 trials (2).

Checkpoint inhibitors (like anti-PD-1/PD-L1 and anti-CTLA-4) used in other cancers (e.g., melanoma) are being actively studied in patients with GBM (Clinical trials.gov). GBM tumors can express PD-L1, which contributes to immune resistance by inducing T-cell apoptosis and promoting an immunosuppressive environment. Recently, the CheckMate-143 trial was presented and showed nivolumab did not improve survival over bevacizumab treatment, in the setting of recurrent GBM, but it did show a manageable safety profile (3). Additionally, other clinical trials using ICIs in malignant gliomas have failed to show a clear benefit (4). An effective immune response likely requires both activation of tumor-infiltrating lymphocytes and recruitment of peripheral lymphocytes into the tumor.

To better identify patients who may respond to ICIs, researchers propose developing peripheral blood-based assays to test T cell responsiveness. This could help select patients more likely to benefit from immunotherapy, improving clinical trial outcomes. Thus, this study was initiated to study the peripheral immune response of patients undergoing ICI therapy compared to their overall survival.

## PATIENTS AND METHODS

### Patient Selection

All patients were enrolled at the Center for Cancer Research of the National Cancer Institute (NCI), Bethesda, Maryland, USA (NCT04817254) and signed informed consent. Eligible patients included those ≥ 18 years old with histologically confirmed newly diagnosed primary glioblastoma or gliosarcoma. They had a complete resection of unifocal disease and completed chemoradiation prior to enrollment. Karnofsky performance score was >70. Patients had adequate organ function including an absolute neutrophil count > 1,500 cells/mm, platelets ≥ 100,000 cells/mm, hemoglobin ≥ 9.0 g/dl, bilirubin < 2 x upper limit of normal (ULN), AST/ALT < 2.5x ULN and creatinine < 1.7 mg/dl or GFR > 30 ml/min. Patients must understand and have the ability to sign a written informed consent. Exclusion criteria included patients with prior placement of Gliadel wafers or treatment with TTF, prior treatment with other investigational agents or prior use of immunotherapy. Patients were excluded if they had a history of allergic reaction to nivolumab, ipilimumab, temozolomide or gadolinium contrast.

### Study Design and Drug Administration

This was a Phase 2, open-label, randomized trial of two different doses of ipilimumab in combination with nivolumab and temozolomide. After confirmation of eligibility, patients were randomized to either Arm 1: nivolumab (1 mg/kg IV every 2 weeks for cycles 1-4, then at a flat dose of 480mg every 4 weeks for cycles 5-16), plus ipilimumab (1 mg/kg IV every 4 weeks for cycles 1-4), plus temozolomide (150 mg/m2 dose on days 1-5 of cycles 1-6), or Arm 2: with the same dosages of nivolumab and temozolomide but a higher dose of ipilimumab (3 mg/kg dose IV every 4 weeks for cycles 1-4). The nivolumab was administered as a 60m IV infusion and was followed by a 60m IV infusion of ipilimumab on day one of each cycle. Temozolomide was taken orally on days 1-5 of the first 6 cycles. Research blood samples were drawn prior to the initiation of treatment of cycle 1 and stored in the laboratory.

### Assessment of T-cell response to activation

A bead-based platform to evaluate T cell response to activation in the presence of inhibitory signals was used. This platform comprises commercially procured tosylactivated Dynabeads conjugated in the laboratory with 50µg of protein, based on an optimized protocol derived from the manufacturer’s instructions. The beads are conjugated with anti-CD3 to provide TCR stimulation, mixed with 100,000 T cells isolated from cryopreserved blood for 4 days, and proliferation measured using flow cytometry +/- the addition of ipilimumab and nivolumab. The instrument software was used to calculate the precursor frequency and expansion index. For the CD4 expansion index, value ≥3 was *reactive*, ≤2 was *suppressed* and 2.1-2.9 was *indeterminant*. For the CD4 precursor frequency, a value ≥40 was *reactive*, ≤30 was *suppressed* and 31-39 was *indeterminant*. For the CD8 expansion index, value ≥4 was *reactive*, ≤3 was *suppressed* and 3.1-3.9 was *indeterminant*. For the CD8 precursor frequency, value ≥50 was *reactive*, ≤40 was *suppressed* and 41-49 was *indeterminant*.

### Statistical Analysis

The primary endpoint was to determine if participants who demonstrate an immune response in peripheral blood T lymphocytes with immune checkpoint inhibitors have an improved outcome compared with those participants who do not demonstrate an immune response as measured by overall survival. Overall survival was defined as the time from treatment initiation to the time of death.

### Data Availability

Raw data for this study were generated at the NCI. Derived data supporting the findings of this study are available from the corresponding author upon request.

## RESULTS/DISCUSSION

Forty patients were treated on this study from 2021-2024. Seventeen of the patients are deceased and had adequate samples for analysis. The OS for these 17 patients was 19m (range 9-45m). Pre-treatment samples for these 17 patients were used to determine the immune reactivity for each patient and whether that reactivity could be used as a biomarker for their OS. The first measurement was CD4 precursor frequency. The range for these values was 4.45-81.42 and 4 patients were classified as *reactive*, 4 as *indeterminate* and 9 as *suppressed*. The OS for the *reactive* patients was 18.5m and 21.6m for the *suppressed* (p=not significant (NS)). For the CD4 expansion index the range of values was from 0.09-4.53 and 2 patients were classified as *reactive*, 3 as *indeterminant* and 12 as *suppressed*. The OS for the *reactive* patients was 17.5m and for the *suppressed* 21m (p=NS). For the CD8 precursor frequency there was a range of values from 11.9-71 and 7 patients were classified as *reactive*, 1 as *indeterminant* and 9 as *suppressed*. The OS for the *reactive* patients was 21.3m and for the *suppressed* 17.6m (p=0.48). The final measurement was the CD8 expansion index and values ranged from 0.38-7.0 and 7 patients were classified as *reactive*, one and *indeterminant* and 9 as *suppressed*. The OS for the *reactive* patients was 20.7m and for the *suppressed* 18.3m (p=NS).

In conclusion, these data do not support the use of CD4 or CD8 precursor frequency or expansion index as biomarkers for ICI treatment efficacy, measured by OS, in patients with GBM. Although the study may have been underpowered to detect meaningful associations, no trend consistent with the hypothesis was observed in this limited group. Notably, only two patients had the same immune classification across all four measurements, making it unclear whether this reflects sampling problem or insufficient sensitivity of the biomarker themselves. Overall, peripheral T-cell reactivity did not correlate with OS in patients receiving ICI therapy.

## Notes

Disclosure of Potential Conflicts of Interest: The authors declare no potential conflicts of interest.

Funding: This work was supported by the Intramural Research Program of the National Cancer Institute at the National Institutes of Health (ZID BC 011642).

### Competing Interest Statement

The authors have declared no competing interest.

### Clinical Trial

NCT04817254

### Funding Statement

This work was supported by the Intramural Research Program of the National Cancer Institute at the National Institutes of Health (ZID BC 011642).

### Author Declarations

All patients were enrolled on a therapy protocol approved by the Institution Review Board of the Center for Cancer Research of the National Cancer Institute (NCI), Bethesda, Maryland, USA (NCT04817254) and signed informed consent.

